# Factors associated with COVID-19 related hospitalisation, critical care admission and mortality using linked primary and secondary care data

**DOI:** 10.1101/2021.01.19.20241844

**Authors:** Lisa Cummins, Irene Ebyarimpa, Nathan Cheetham, Victoria Tzortziou Brown, Katie Brennan, Jasmina Panovska-Griffiths

## Abstract

**Background:** To identify risk factors associated with increased risk of hospitalisation, intensive care unit (ICU) admission and mortality in inner North East London (NEL) during the first UK COVID-19 wave.

**Methods:** Multivariate logistic regression analysis on linked primary and secondary care data from people aged 16 or older with confirmed COVID-19 infection between 01/02/2020-30/06/2020 determined odds ratios (OR), 95% confidence intervals (CI) and p-values for the association between demographic, deprivation and clinical factors with COVID-19 hospitalisation, ICU admission and mortality.

**Results:** Over the study period 1,781 people were diagnosed with COVID-19, of whom 1,195 (67%) were hospitalised, 152 (9%) admitted to ICU and 400 (23%) died. Results confirm previously identified risk factors: being male, or of Black or Asian ethnicity, or aged over 50. Obesity, type 2 diabetes and chronic kidney disease (CKD) increased the risk of hospitalisation. Obesity increased the risk of being admitted to ICU. Underlying CKD, stroke and dementia increased the risk of death. Having learning disabilities was strongly associated with increased risk of death (OR=4.75, 95%CI=(1.91,11.84), p=0.001). Having three or four co-morbidities increased the risk of hospitalisation (OR=2.34,95%CI=(1.55,3.54),p<0.001;OR=2.40, 95%CI=(1.55,3.73), p<0.001 respectively) and death (OR=2.61, 95%CI=(1.59,4.28), p<0.001;OR=4.07, 95% CI= (2.48,6.69), p<0.001 respectively).

**Conclusions:** We confirm that age, sex, ethnicity, obesity, CKD and diabetes are important determinants of risk of COVID-19 hospitalisation or death. For the first time, we also identify people with learning disabilities and multi-morbidity as additional patient cohorts that need to be actively protected during COVID-19 waves.

## Introduction

The COVID-19 pandemic caused by the Severe Acute Respiratory Syndrome – Coronavirus 2 (SARS-CoV-2) continues to spread worldwide with over 48 million cases and over 1.22 million deaths reported worldwide as 5 November 2020 (1). In the UK, the first two reported cases were on 31 January 2020 and the first reported COVID-19 related death was on 6 March 2020. As of 5 November 2020, 1.1 million people tested positive for COVID-19 and, of those, 47,742 died within 28 days of a positive test (2).

Existing studies suggest that people of Asian or Black ethnicity, those over the age of 70 and males have a significantly higher risk of COVID-19 related mortality (3,4). Also, areas with higher levels of deprivation, higher proportions of people from Asian or Black ethnic backgrounds or higher proportions of people with pre-existing health conditions tend to be more adversely affected by COVID-19 (5–7). The Chinese Center for Disease Control and Prevention reported, in a study of 44,672 individuals with 1,023 deaths, that cardiovascular disease, hypertension, diabetes, respiratory disease and cancers were associated with an increased risk of death from COVID-19 (8). A UK cross-sectional survey of 16,749 people who were hospitalised with COVID-19 showed the risk of death was higher in people with underlying cariac, pulmonary and kidney disease, as well as cancer, dementia and obesity (9). Obesity was also associated with higher risk of COVID-19 hospitalisation in New York (10).

North East London (NEL) has a diverse population, with high levels of deprivation and inequality. Residents in Newham, Hackney, Tower Hamlets, and Barking and Dagenham are comparatively younger (11) and around half of all residents are from Black, Asian and Minority Ethnic (BAME) communities; ranging from around 16% in Havering to 68% in Newham (12). In terms of deprivation, Barking and Dagenham, Newham, Tower Hamlets, and Hackney are among the most deprived areas in England, and inner NEL wards tend to be more deprived but pockets of deprivation persist in outer boroughs also (13). The difference between the lowest and highest healthy life expectancy across the boroughs is 9.3 and 8.0 years for men and women, respectively (14).

Boroughs in North East London (NEL) have been disproportionally affected by the COVID-19 epidemic, with Newham (203.4) and Hackney (186.8) having the second and third highest COVID-19 age-standardised mortality rate per 100,000 population in the UK (see Figure **1**) (6). The first case of COVID-19 in the region was reported on 19 February 2020, and the first COVID-19 associated death was documented on 6 March 2020 (2). An estimated 970,000 people live in the North East London areas of City and Hackney, Newham and Tower Hamlets (11) and, as of 4 November 2020, there have been 11,886 confirmed cases of COVID-19 in the area (2).

On March 23, 2020, the UK Government imposed strict social distancing measures (‘lock-down’), to protect the public, slow down the virus spread, reduce the associated morbidity and mortality, and prevent excess demand on the National Health Service (NHS). In early June 2020, measures were gradually eased with the partial reopening of primary schools and secondary schools before non-essential businesses reopened on 4 July 2020. Following a period of slow growth in cases during July and August, cases started to rise again in September 2020. In response, the UK Government introduced a Tiered system of local lockdowns; however, on 5 November 2020, a second national lockdown was imposed following a sustained increase in cases and a rise in hospitalisations (2,15).

Analysing data on the local population is integral to the work of the North East London Integrated Health and Care System to ensure the development of equitable and appropriate epidemic response plans for managing future waves of COVID-19 as well as supporting the system and individuals recovering from COVID-19. Preliminary findings of this work have already been used to support system planning, in particular, information on cohorts at greater risk of infection and hospitalisation has informed the public health response locally.

In this paper, we determined which population cohorts are at increased risk of hospitalisation, ICU admission and death following a diagnosis of COVID-19 in the NEL areas of City of London and Hackney, Newham and Tower Hamlets. To achieve this, we used local health care data on demographic characteristics and clinical presentations of 1,781 people (registered with a GP practice in the areas) with a confirmed diagnosis of COVID-19 between 1 February 2020 and 30 June 2020.

Our overall objective was to focus on an ethnically diverse area with high levels of deprivation. We feel this is crucial for understanding the impact of demographic and health factors on COVID-19-related hospitalisation and mortality as well as identifying the common clinical conditions experienced by people living in an urban setting who are at risk of adverse outcomes related to COVID-19. Our findings are broad and translatable to other settings.

## Methods

### Data sources and observation period

Unique patient identifiers were used to obtain service use and mortality outcome data from the Secondary Uses Service (SUS) hospital inpatient data (16). Between 1 February 2020 and 30 June 2020, 1,781 people aged 16 or older (registered with a GP practice in Newham, Tower Hamlets and City and Hackney) were recorded as being COVID-19 positive in the primary or secondary care dataset. Data on sociodemographic factors (gender, age, self-reported ethnicity, and English indices of deprivation (IMD) 2019 score (13)), smoking status and obesity were extracted from GP data along with mortality data where recorded. Data on 17 clinical conditions, primarily long-term conditions (LTCs), included in the UK Quality and Outcomes Framework (QOF) were also extracted from GP data (17).

### Statistical analysis

Multivariate logistic regression was used to estimate the association between demographic, socioeconomic and clinical factors of people with confirmed COVID-19 and hospitalisation, admission to ICU or COVID-19-related mortality. The analysis was done in R programming language.

Demographic covariates included gender, age, stratified into 16-49, 50-69 and 70+ years of age, and self-reported ethnicity grouped into White, Black, Asian, Mixed, Other, and unknown. Given that 92% of people included in the analysis were living in neighbourhoods that are within the 40% most deprived areas in England, a binary socioeconomic covariate to indicate if people were living in the 30% most deprived areas was used in the statistical analysis rather than stratifying into quintiles. Smoking status and obesity were also included as covariates.

The clinical covariates comprised 17 clinical factors included in the UK Quality and Outcomes Framework (17): asthma; atrial fibrillation; cancer; chronic heart disease (CHD); chronic kidney disease (CKD); chronic obstructive pulmonary disease (COPD); dementia; depression; diabetes (Type 1 and Type 2 diabetes); epilepsy; heart failure; hypertension; learning disability; severe mental illness; peripheral arterial disease (PAD); and stroke.

The association between the covariates and the three outcome variables (hospitalisation, ICU admission and death following COVID-19 infection) were examined using two separate multivariate logistic regression models. The first model included the demographic and socioeconomic factors as well as obesity, smoking status and the 17 individual clinical factors as covariates. The second model replaced the 17 individual clinical covariates with a measure of multiple morbidities defined as counts of clinical factors per person. The counts were stratified into five categories, ranging from none to four or more clinical factors. The odds ratios (OR) and their 95% confidence intervals (95% CI) as well as the p-value for the associations within the two models were derived.

### Role of the funding source

The funders had no role in the study design, data collection, analysis or interpretation of the data or writing of the report. The corresponding author had access to aggregate data and the final responsibility to submit for publication. LC, IE and NC had full access to all of the data.

## Results

### Descriptive statistics results

Over the study period 1,781 people aged 16 or older (registered with a GP practice in Newham, Tower Hamlets and City and Hackney) were identified as being COVID-19 positive across the primary and secondary care datasets.

Of those with a confirmed diagnosis of COVID-19, 1,195 (67%) were hospitalised, 152 (9%) were admitted to ICU, and 400 (23%) died. Over two-thirds (68%) were aged 50 or older and of those 76% were hospitalised, 10% admitted to ICU and 31% died. Around 60% of COVID-19 confirmed cases had ethnicity recorded as Black, Asian or Mixed and of these 69% were hospitalised, 10% admitted to ICU and 22% died. 45% of the cohort was female and almost all (92%) people were living in neighbourhoods that are within the 40% most deprived areas in England. A full summary of descriptive statistics is shown in Table 1.

**Table 1:** Results of the descriptive statistics.

| Category/characteristic | Number of people diagnosed with Covid-19 |  |  |  |
| --- | --- | --- | --- | --- |
|  | Total | Hospitalisations | ICU admission | Deaths |
| All | N = 1,781<br>n (%) | N = 1,195<br>n (%) | N = 152<br>n (%) | N = 400<br>n (%) |
| **Gender** |  |  |  |  |
| Female | 797 (44.8%) | 454 (38.0%) | 36 (23.7%) | 132 (33.0%) |
| Male | 984 (55.2%) | 741 (62.0%) | 116 (76.3%) | 268 (67.0%) |
| **Age group** |  |  |  |  |
| 16-49 | 563 (31.6%) | 274 (22.9%) | 32 (21.1%) | 20 (5.0%) |
| 50-69 | 637 (35.8%) | 467 (39.1%) | 96 (63.2%) | 124 (31.0%) |
| 70+ | 581 (32.6%) | 454 (38.0%) | 24 (15.8%) | 256 (64.0%) |
| **IMD quintile** |  |  |  |  |
| Least deprived | 678 (38.1%) | 454 (38.0%) | 57 (37.5%) | 151 (37.8%) |
| Second Least deprived | 952 (53.5%) | 654 (54.7%) | 79 (52.0%) | 213 (53.2%) |
| Third Least deprived | 109 (6.1%) | 62 (5.2%) | 11 (7.2%) | 27 (6.8%) |
| Second most deprived | 31 (1.7%) | 19 (1.6%) | 5 (3.3%) | 6 (1.5%) |
| Most deprived | 11 (0.6%) | 6 (0.5%) | 0 (0.0%) | 3 (0.8%) |
| Living in the 30% most deprived | 1392 (78.2%) | 944 (79.0%) | 116 (76.3%) | 310 (77.5%) |
| **Ethnic Group** |  |  |  |  |
| White | 626 (35.1%) | 405 (33.9%) | 37 (24.3%) | 144 (36.0%) |
| Asian | 606 (34.0%) | 396 (33.1%) | 67 (44.1%) | 125 (31.2%) |
| Black | 425 (23.9%) | 315 (26.4%) | 36 (23.7%) | 108 (27.0%) |
| Mixed | 33 (1.9%) | 20 (1.7%) | 0 (0.0%) | 6 (1.5%) |
| Other | 81 (4.5%) | 54 (4.5%) | 11 (7.2%) | 14 (3.5%) |
| Not known or Not stated | 10 (0.6%) | 5 (0.4%) | 1 (0.7%) | #N/A |
| Obese | 482 (27.1%) | 366 (30.6%) | 58 (38.2%) | 115 (28.8%) |
| Smoker (current) | 181 (10.2%) | 104 (8.7%) | 9 (5.9%) | 31 (7.8%) |
| Asthma | 244 (13.7%) | 166 (13.9%) | 14 (9.2%) | 55 (13.8%) |
| Atrial fibrillation | 107 (6.0%) | 82 (6.9%) | 3 (2.0%) | 49 (12.2%) |
| Cancer | 148 (8.3%) | 118 (9.9%) | 13 (8.6%) | 60 (15.0%) |
| Chronic heart disease | 217 (12.2%) | 170 (14.2%) | 20 (13.2%) | 94 (23.5%) |
| Chronic kidney disease | 365 (20.5%) | 297 (24.9%) | 28 (18.4%) | 165 (41.2%) |
| Chronic obstructive pulmonary disease | 145 (8.1%) | 114 (9.5%) | 6 (3.9%) | 57 (14.2%) |
| Dementia | 119 (6.7%) | 83 (6.9%) | 1 (0.7%) | 63 (15.8%) |
| Depression | 216 (12.1%) | 136 (11.4%) | 14 (9.2%) | 36 (9.0%) |
| Type 1 diabetes | 16 (0.9%) | 10 (0.8%) | 2 (1.3%) | 4 (1.0%) |
| Type 2 diabetes | 625 (35.1%) | 497 (41.6%) | 66 (43.4%) |  |
| Epilepsy | 35 (2.0%) | 27 (2.3%) | 2 (1.3%) | 13 (3.2%) |
| Heart failure | 139 (7.8%) | 110 (9.2%) | 4 (2.6%) | 61 (15.2%) |
| Hypertension | 825 (46.3%) | 630 (52.7%) | 81 (53.3%) | 273 (68.2%) |
| Learning disability | 28 (1.6%) | 22 (1.8%) | 2 (1.3%) | 11 (2.8%) |
| Severe mental illness | 85 (4.8%) | 62 (5.2%) | 8 (5.3%) | 18 (4.5%) |
| Peripheral arterial disease | 55 (3.1%) | 43 (3.6%) | 3 (2.0%) | 26 (6.5%) |
| Stroke | 138 (7.7%) | 104 (8.7%) | 6 (3.9%) | 68 (17.0%) |
| Hospitalised | 1195 (67.1%) | - | 152 (100.0%) | 363 (90.8%) |
| Critical care | 152 (8.5%) | 152 (12.7%) | - | 78 (19.5%) |
| Died | 400 (22.5%) | 363 (30.4%) | 78 (51.3%) | - |

### Regression analysis results

The associations between patient-level factors and risk of COVID-19-related hospitalisations, admission to ICUs and COVID-19 related death are shown in Tables 2-3, with the ORs for COVID-19 hospitalisation, ICU admission and death plotted in Figures 2-4.

**Table 2:** Results of multivariate logistic regression analysis that estimates the association between demographic and socioeconomic factors as well as obesity, smoking status and 17 individual clinical factors and the three outcome variables (hospitalisation, ICU admission and death following COVID-19 infection).^*^p<0.1;^*,*^p<0.05;^*,*,*^p<0.01

| Dependent variables | Hospitalisations | ICU admission | Deaths |
| --- | --- | --- | --- |
| Independent variables | Odds Ratios (95% confidence interval) and p-value |  |  |
| **Gender**<br>(reference: female) |  |  |  |
| Male | 2.39*** (1.92 - 2.97)<br>p < 0.001 | 2.78*** (1.85 - 4.18)<br>p < 0.001 | 1.91*** (1.46 - 2.51)<br>p < 0.001 |
| **Age**<br>(reference: 16-49) |  |  |  |
| 50-69 | 2.23*** (1.70 - 2.94)<br>p < 0.001 | 2.57*** (1.61 - 4.11)<br>P < 0.001 | 5.33*** (3.15 - 9.01)<br>p < 0.001 |
| 70+ | 2.97*** (2.05 - 4.30)<br>p < 0.001 | 0.86 (0.44 - 1.68)<br>p = 0.660 | 12.65*** (7.22 - 22.17)<br>p = 0.000 |
| **Ethnicity**<br>(reference: White) |  |  |  |
| Asian | 1.28* (0.97 - 1.69)<br>p = 0.076 | 1.62** (1.01 - 2.59)<br>p = 0.045 | 1.71*** (1.21 - 2.42)<br>p = 0.002 |
| Black | 1.54*** (1.13 - 2.09)<br>p = 0.006 | 1.19 (0.70 - 2.00)<br>p = 0.523 | 1.37* (0.96 - 1.94)<br>p = 0.084 |
| Mixed | 1.30 (0.59 - 2.85)<br>p = 0.512 | 0.00 (0.00 - Inf)<br>p = 0.972 | 1.55 (0.55 - 4.38)<br>p = 0.409 |
| Other | 1.38 (0.81 - 2.35)<br>p = 0.240 | 1.77 (0.82 - 3.81)<br>p = 0.147 | 1.45 (0.72 - 2.92)<br>p = 0.296 |
| Not known | 0.99 (0.26 - 3.86)<br>p = 0.993 | 1.68 (0.18 - 15.73)<br>p = 0.648 | 7.00** (1.42 - 34.37)<br>p = 0.017 |
| **Top 30% most deprived areas** | 1.12 (0.86 - 1.45)<br>p = 0.388 | 0.85 (0.56 - 1.30)<br>p = 0.462 | 0.89 (0.65 - 1.21)<br>p = 0.453 |
| **Obese** | 1.64*** (1.25 - 2.15)<br>p < 0.001 | 1.74*** (1.18 - 2.56)<br>p = 0.005 | 1.15 (0.86 - 1.55)<br>p = 0.355 |
| **Smoker (current)** | 0.63*** (0.44 - 0.88)<br>p = 0.008 | 0.60 (0.29 - 1.22)<br>p = 0.156 | 0.78 (0.49 - 1.24)<br>p = 0.296 |
| **Asthma** | 1.05 (0.77 - 1.45)<br>p = 0.741 | 0.68 (0.38 - 1.24)<br>p = 0.209 | 1.03 (0.70 - 1.50)<br>p = 0.883 |
| **Atrial fibrillation** | 0.88 (0.52 - 1.48)<br>p = 0.622 | 0.51 (0.15 - 1.75)<br>p = 0.285 | 1.19 (0.74 - 1.92)<br>p = 0.465 |
| **Cancer** | 1.39 (0.88 - 2.17)<br>p = 0.155 | 1.30 (0.68 - 2.48)<br>p = 0.432 | 1.46* (0.98 - 2.17)<br>p = 0.063 |
| **CHD** | 0.86 (0.58 - 1.27)<br>p = 0.446 | 1.17 (0.66 - 2.08)<br>p = 0.590 | 1.14 (0.80 - 1.63)<br>p = 0.454 |
| **CKD** | 1.55** (1.10 - 2.19)<br>p = 0.012 | 1.22 (0.73 - 2.04)<br>p = 0.443 | 1.74*** (1.28 - 2.36)<br>p < 0.001 |
| **COPD** | 1.35 (0.85 - 2.15)<br>p = 0.209 | 0.65 (0.27 - 1.60)<br>p = 0.351 | 1.11 (0.73 - 1.69)<br>p = 0.632 |
| **Dementia** | 0.67* (0.42 - 1.07)<br>p = 0.094 | 0.16* (0.02 - 1.22)<br>p = 0.077 | 2.16*** (1.40 - 3.33)<br>p = 0.001 |
| **Depression** | 0.98 (0.71 - 1.36)<br>p = 0.905 | 0.85 (0.46 - 1.57)<br>p = 0.612 | 0.92 (0.60 - 1.41)<br>p = 0.702 |
| **Type 1 diabetes** | 0.88 (0.29 - 2.67) | 1.94 (0.39 - 9.56) | 1.53 (0.39 - 5.93) |
|  | p = 0.815 | p = 0.414 | p = 0.539 |
| **Type 2 diabetes** | 1.53*** (1.17 - 2.00)<br>p = 0.002 | 1.01 (0.68 - 1.50)<br>p = 0.966 | 1.23 (0.93 - 1.63)<br>p = 0.152 |
| **Epilepsy** | 1.51 (0.63 - 3.62)<br>p = 0.353 | 0.81 (0.17 - 3.85)<br>p = 0.793 | 1.63 (0.73 - 3.62)<br>p = 0.235 |
| **Heart failure** | 1.06 (0.65 - 1.72)<br>p = 0.829 | 0.34* (0.12 - 1.01)<br>p = 0.051 | 1.09 (0.71 - 1.68)<br>p = 0.685 |
| **Hypertension** | 1.05 (0.80 - 1.37)<br>p = 0.741 | 1.29 (0.86 - 1.95)<br>p = 0.224 | 1.16 (0.86 - 1.56)<br>p = 0.339 |
| **Learning disability** | 2.07 (0.78 - 5.45)<br>p = 0.142 | 1.22 (0.26 - 5.79)<br>p = 0.801 | 4.75*** (1.91 - 11.84)<br>p = 0.001 |
| **Severe mental illness** | 1.04 (0.61 - 1.78)<br>p = 0.873 | 1.42 (0.62 - 3.22)<br>p = 0.405 | 0.72 (0.39 - 1.30)<br>p = 0.274 |
| **Peripheral arterial disease** | 0.94 (0.46 - 1.89)<br>p = 0.859 | 0.83 (0.24 - 2.89)<br>p = 0.768 | 1.37 (0.74 - 2.52)<br>p = 0.316 |
| **Stroke** | 0.88 (0.56 - 1.36)<br>p = 0.560 | 0.52 (0.21 - 1.25)<br>p = 0.144 | 1.93*** (1.29 - 2.88)<br>p = 0.001 |

**Figure 1:**
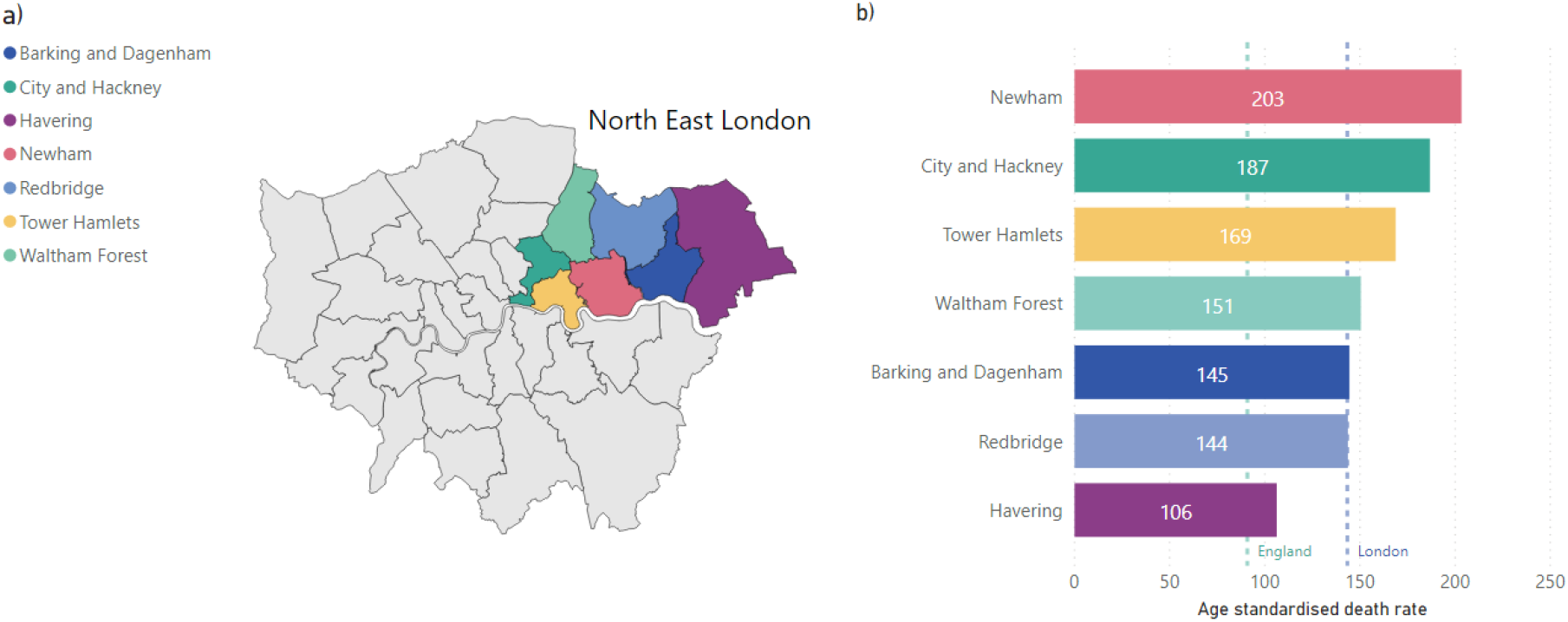
a) NHS Clinical Commissioning Group (CCG) London boundaries, with North East London CCGs highlighted. b) Age-standardised COVID-19 mortality rates (deaths occurring between 1 March and 31 July 2020) by North East London boroughs(6).

**Figure 2:**
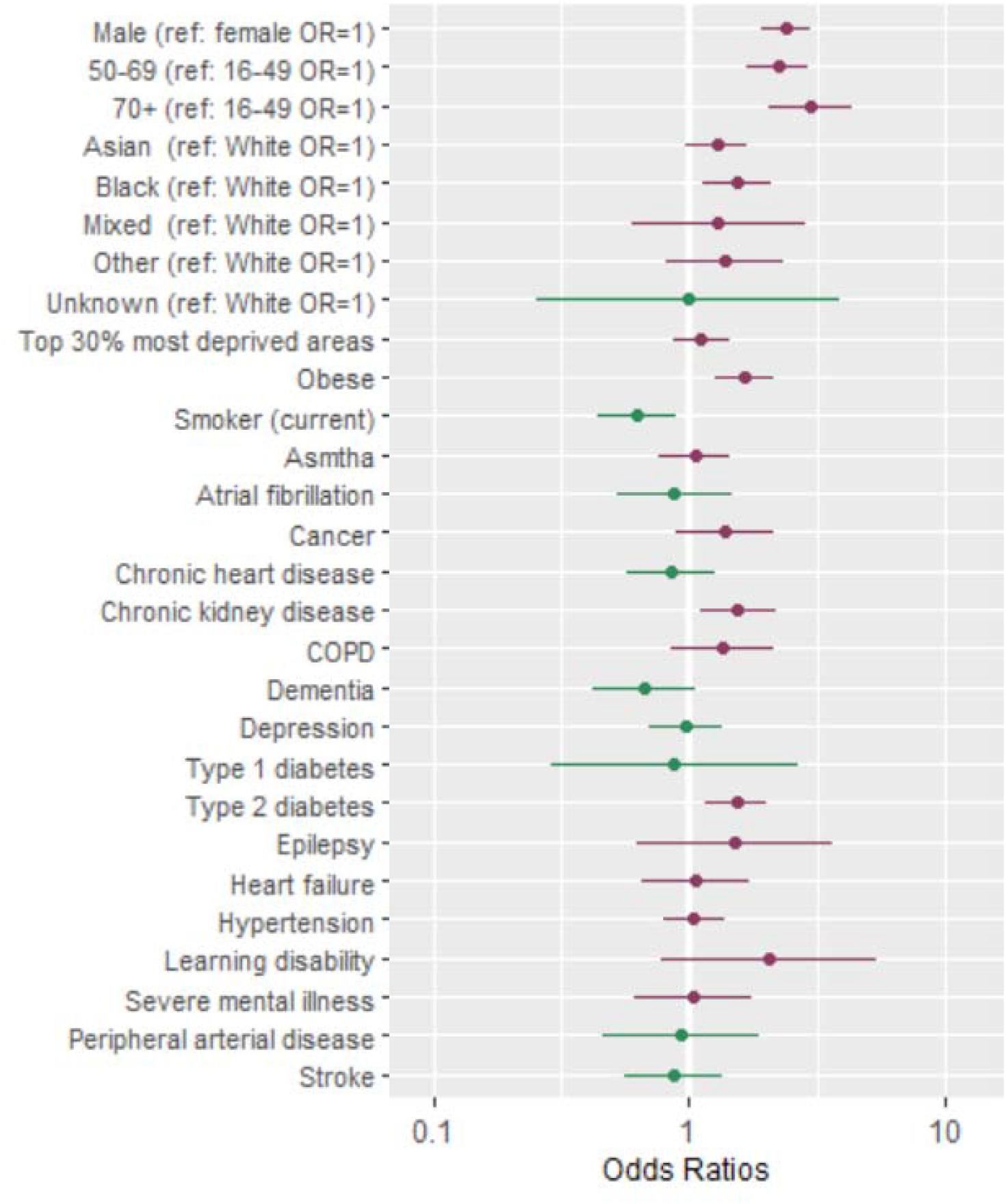
Forest plot showing odds ratios (OR) of hospitalisation following a COVID-19 diagnosis. Dots represent OR shown on a log scale while the error bars represent the limits of the 95% confidence intervals. ref = reference group. Table 2, column 2 contains the exact ORs and p-values of the association.

#### Gender, age, ethnicity and deprivation

Our results indicate that being male, compared to being female, was associated with increased risk of COVID-19 hospitalisation (OR=2.39, 95%CI=(1.92, 2.97), p<0.001), ICU admission (OR=2.78, 95% CI=(1.85, 4.18), p<0.001) and death (OR=1.91, 95%CI=(1.46, 2.51), p<0.001) (Table 2).

Compared to younger people (aged between 16 and 49), people aged over 50 were at higher risk of being hospitalised or dying following a diagnosis of COVID-19. People aged 70 or older had a much greater likelihood of hospitalisation (OR=2.97, 95%CI=(2.05, 4.30), p<0.001) and death (OR= 12.65, 95%CI= (7.22, 22.17), p<0.001) while people aged 50-69 were more likely to be hospitalised (OR 2.23, 95% CI=(1.70, 2.94), p<0.001), admitted to ICU (OR=2.57, 95%CI=(1.61, 4.11), p<0.001) and die (OR=5.33, 95% CI=(3.15, 9.01), p< 0.001) compared to those aged 16-49 (Table 2).

Compared to people of White ethnicity, people of Black ethnicity were at higher risk of COVID-19-associated hospitalisation (OR=1.54, 95%CI=(1.13, 2.09), p=0.006), while people of Asian ethnicity were at higher risk of ICU admission (OR=1.62, 95%CI=(1.01, 2.59), p=0.045) and COVID-19 associated death (OR=1.71, 95%CI=(1.21, 2.42), p=0.002) (Table 2 and Figure 2).

The effect of deprivation was small and not statistically significant for all outcome measures (Tables 2); however, this is likely due to the majority of people (92%) living in the top 40% most deprived areas.

#### Multi-morbidity

Our results indicate that people with multi-morbidity are at increased risk of being hospitalised or dying following COVID-19 infection (Table 3), with the odds increasing as the number of underlying clinical conditions increases. Having three co-morbidities was associated with increased risk of COVID-19 hospitalisation (OR=2.34, 95% CI=(1.55, 3.54), p < 0.001), and death (OR=2.61, 95% CI=(1.59, 4.28), p < 0.001). The odds of admission to ICU was weaker and not statistically significant (OR=1.75, 95% CI=(0.97, 3.14), p = 0.062). Having four or more co-morbidities was associated with even greater risk of death (OR=4.07, 95%CI=(2.48, 6.69), p<0.001) and with increased hospitalisation (OR=1.40, 95%CI=(1.55, 3.73), p<0.001) but not with increased admission to ICU (OR=0.92, 95%CI=(0.46, 1.82), p=0.803).

**Table 3:** Results of multivariate logistic analysis that estimates the association between demographic and socioeconomic factors as well as obesity, smoking status and counts of clinical factors per person and the three outcome variables (hospitalisation, ICU admission and death following COVID-19 infection). In comparison to Table 2, the regression model that replaced the 17 individual clinical covariates with a measure of multiple morbidities defined as counts of clinical factors per person. The counts were stratified into five categories, ranging from none to four or more clinical factors. ^*^p<0.1;^*,*^p<0.05;^*,*,*^p<0.01

| Dependent variables | Hospitalisations | ICU admission | Deaths |
| --- | --- | --- | --- |
| Independent variables | Odds Ratios (95% confidence interval) and p-value |  |  |
| **Gender**<br>(reference: female) |  |  |  |
| Male | 2.37*** (1.91 - 2.94)<br>p < 0.001 | 2.82*** (1.89 - 4.21)<br>p < 0.001 | 1.88*** (1.45 - 2.45)<br>p < 0.001 |
| **Age**<br>(reference: 16-49) |  |  |  |
| 50-69 | 2.13*** (1.63 - 2.79)<br>p < 0.001 | 2.51*** (1.58 - 3.97)<br>p < 0.001 | 4.57*** (2.74 - 7.64)<br>p < 0.001 |
| 70+ | 2.63*** (1.90 - 3.65)<br>p < 0.001 | 0.65 (0.35 - 1.23)<br>p = 0.187 | 13.67*** (8.05 - 23.22)<br>p < 0.001 |
| **Ethnicity**<br>(reference: White) |  |  |  |
| Asian | 1.32** (1.01 - 1.72)<br>p = 0.040 | 1.74** (1.11 - 2.71)<br>p = 0.015 | 1.57*** (1.14 - 2.17)<br>p = 0.006 |
| Black | 1.54*** (1.15 - 2.07)<br>p = 0.004 | 1.28 (0.78 - 2.11)<br>p = 0.327 | 1.29 (0.93 - 1.79)<br>p = 0.121 |
| Mixed | 1.29 (0.59 - 2.82)<br>p = 0.528 | 0.00 (0.00 - Inf)<br>p = 0.973 | 1.38 (0.50 - 3.82)<br>p = 0.540 |
| Other | 1.43 (0.84 - 2.44)<br>p = 0.183 | 2.06* (0.96 - 4.39)<br>p = 0.062 | 1.31 (0.66 - 2.62)<br>p = 0.439 |
| **Not known** | 1.02 (0.26 - 3.98)<br>p = 0.978 | 1.82 (0.19 - 17.87)<br>p = 0.607 | 6.38** (1.24 - 32.77)<br>p = 0.026 |
| **Top 30% most deprived areas** | 1.12 (0.86 - 1.44)<br>p = 0.397 | 0.85 (0.56 - 1.29)<br>p = 0.447 | 0.95 (0.70 - 1.29)<br>p = 0.742 |
| **Obese** | 1.58*** (1.22 - 2.06)<br>p = 0.001 | 1.68*** (1.15 - 2.46)<br>p = 0.008 | 0.95 (0.71 - 1.26)<br>p = 0.707 |
| **Smoker (current)** | 0.63*** (0.45 - 0.88)<br>p = 0.007 | 0.55 (0.27 - 1.12)<br>p = 0.100 | 0.72 (0.46 - 1.13)<br>p = 0.149 |
| **Multi-morbidity**<br>(reference: no co-morbidities) |  |  |  |
| 1 co-morbidity | 1.29* (0.96 - 1.73)<br>p = 0.090 | 1.09 (0.65 - 1.82)<br>p = 0.755 | 1.48 (0.91 - 2.39)<br>p = 0.113 |
| 2 co-morbidities | 1.40** (1.01 - 1.94)<br>p = 0.043 | 1.01 (0.59 - 1.75)<br>p = 0.960 | 2.55*** (1.60 - 4.04)<br>p = 0.000 |
| 3 co-morbidities | 2.34*** (1.55 - 3.54)<br>p < 0.001 | 1.75* (0.97 - 3.14)<br>p = 0.062 | 2.61*** (1.59 - 4.28)<br>p < 0.001 |
| 4+ co-morbidities | 2.40*** (1.55 - 3.73)<br>p < 0.001 | 0.92 (0.46 - 1.82)<br>p = 0.803 | 4.07*** (2.48 - 6.69)<br>p < 0.001 |

#### Clinical factors associated with all outcomes

A range of clinical factors and conditions were determined to be associated with COVID-19-related hospitalisation (Table 2 and Figure 2)), admission to ICU (Table 2 and Figure 3) and deaths (Table 2 and Figure 4). The key clinical risk factors for hospitalisation with COVID-19 were being obese (OR=1.64, 95%CI=(1.25, 2.15), p=0.000) or having underlying CKD (OR=1.55, 95%CI=(1.10, 2.19), p=0.012) or Type 2 diabetes (OR=1.53, 95%CI=(1.17, 2.00), p=0.002). The key clinical risk factor for admission to ICU with COVID-19 was being obese (OR=1.74, 95%CI=(1.18, 2.56), p=0.005). The key clinical risk factors for dying from COVID-19 were having underlying CKD (OR=1.74, 95%CI=(1.28, 2.36), p=0.000) or learning disability (OR=4.75, 95%CI=(1.91, 11.84), p=0.001), having dementia (OR=2.16, 95%CI=(1.40, 3.33), p=0.001) or having suffered a stroke (OR=1.93, 95%CI=(1.29, 2.88), p=0.001).

**Figure 3:**
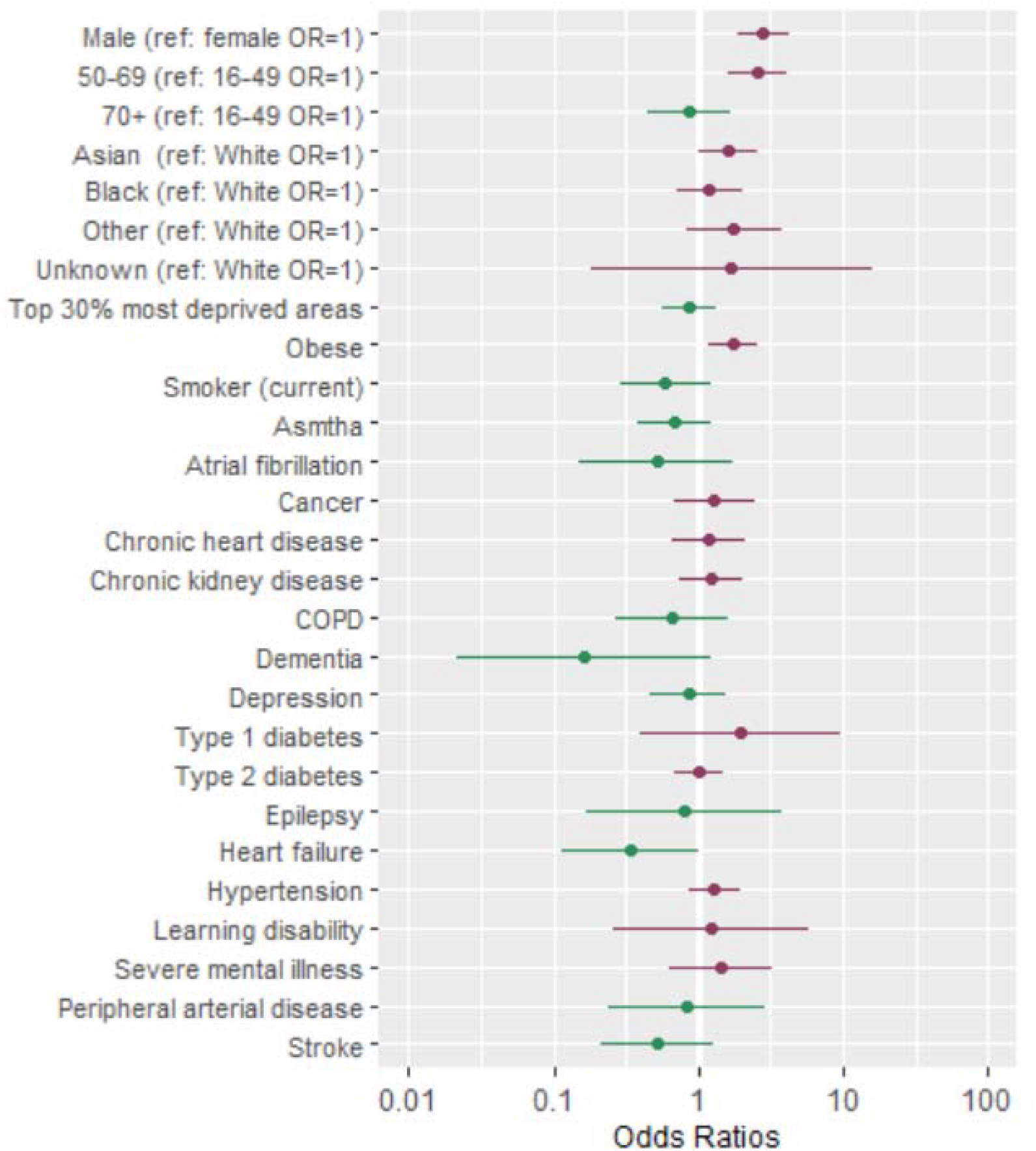
Forest plot showing odds ratios (OR) of admission to critical care following a diagnosis of COVID-19. Dots represent OR shown on a log scale while the error bars represent the limits of the 95% confidence intervals. ref = reference group. Table 2, column 3 contains the exact ORs and p-values of the association.

**Figure 4:**
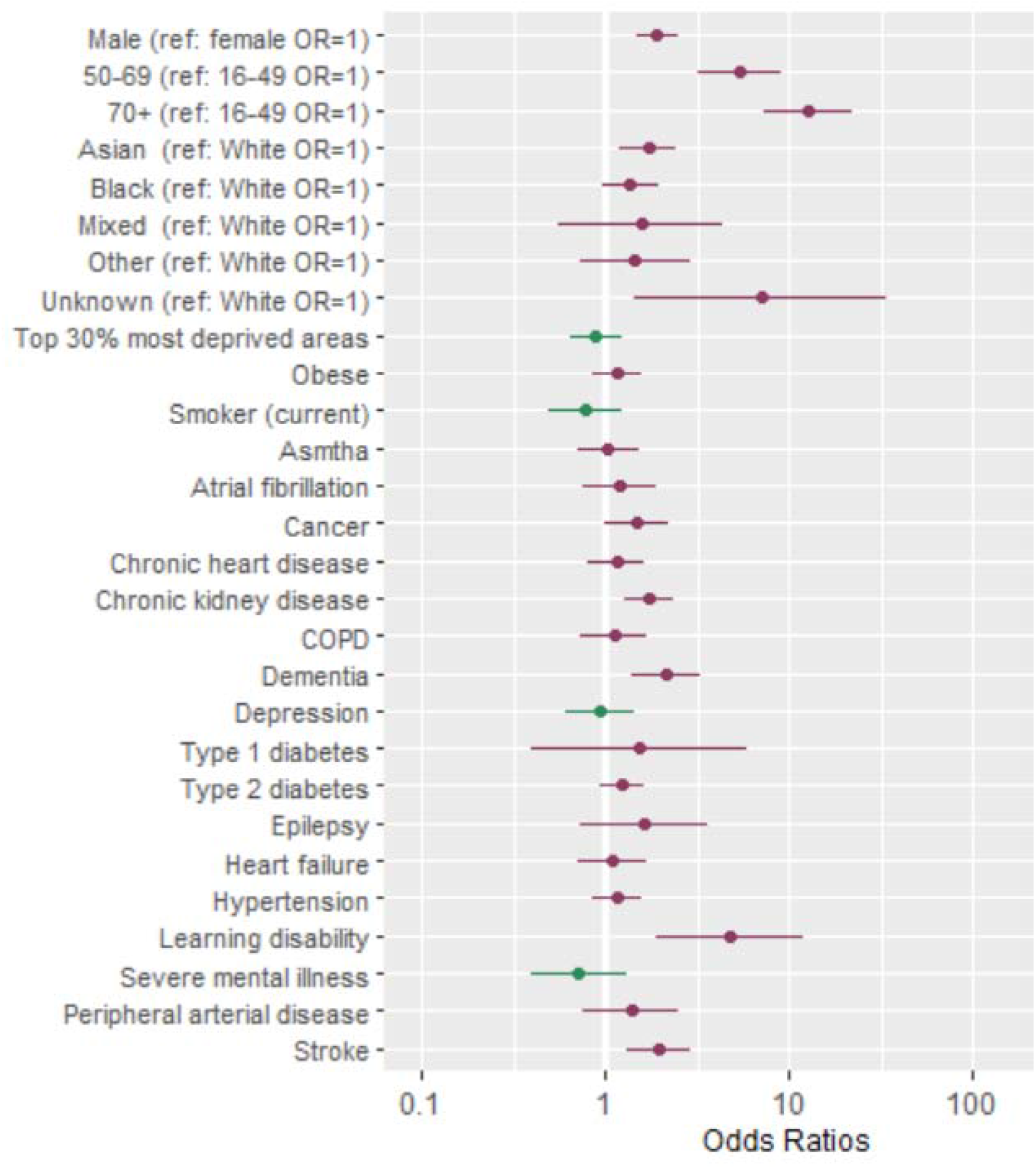
Forest plot showing odds ratios (OR) of dying following a diagnosis of COVID-19. Dots represent OR shown on a log scale while the error bars represent the limits of the 95% confidence intervals. ref = reference group. Table 3, column 4 contains the exact ORs and p-values of the association.

Considering clinical factors separately having CKD was associated with increased risk of COVID-19 related hospitalisation (OR=1.55, 95%CI=(1.10, 2.19), p=0.012) and mortality (OR=1.74, 95%CI=(1.28, 2.36, p=0.000), whereas the odds of admission to ICU was less strong and not statistically significant (OR=1.22, 95%CI=(0.73, 2.04), p=0.443).

People with Type 2 diabetes were also more likely to be hospitalised (OR=1.53, 95%CI=(1.17, 2.00), p=0.002) following a diagnosis with COVID-19, compared to those without; however, the odds of being admitted to ICU (OR=1.01, 95%CI=(0.68, 1.50), p=0.966) or dying (OR=1.23, 95%CI=(0.93, 1.63), p=0.152) were small and not statistically significant.

Being obese was associated with an increased risk of hospitalisation (OR=1.64, 95%CI=(1.25, 2.15), p<0.001) and admission to ICU (OR=1.74, 95%CI=(1.18, 2.56), p=0.005) but not of dying (OR=1.15, 95%CI=(0.86, 1.55), p=0.355), compared to those who were not obese.

People with learning disabilities had the greatest risk of death associated with a single clinical factor following a diagnosis of COVID-19, with an odds ratio of 4.75 (95%CI=1.91, 11.84, p=0.001). This group had a greater, although not statistically significant, risk of being admitted to hospital or ICU.

People who had previously experienced a stroke or had been diagnosed with dementia had an increased risk of COVID-19 related mortality (OR=1.93, 95%CI=(1.29, 2.88), p=0.001 for previous stroke and OR=2.16, 95%CI=(1.40, 3.33), p=0.001 for dementia); however, both groups were less likely be admitted to hospital or ICU.

## Discussion

Our study is the first to use local linked primary and secondary data to examine the key risk factors for COVID-19-related hospitalisation, ICU admission and mortality in an ethnically diverse inner-city area with high levels of deprivation. As well as confirming existing findings that elderly people, people of Asian or Black ethnicity, and those with pre-existing health conditions are at increased risk of COVID-19-related hospitalisation and mortality, our analysis identifies new population cohorts at increased risk of COVID-19 hospitalisation, critical care admission and death.

In line with previous research, being male, Black or Asian ethnicity (across outcomes) or of older age (50 years and over) were found to be associated with COVID-19 related hospitalisation and mortality. In addition, having three or more co-morbidities increased the risk of hospitalisation and death but not ICU admission. Following age, having four or more co-morbidities was the single largest and most reliable patient characteristic associated with higher risk of death (OR=4.07, (95%CI=2.48, 6.69), p=0.000) following a diagnosis of COVID-19, demonstrating the strong compounding effect of medical complexity and multi-morbidity on COVID-19 outcomes.

Importantly, we identified new population cohorts at risk from COVID-19. In particular, for the first time, people with learning disabilities were determined to have five times the odds of dying following a diagnosis of COVID-19 compared to people without.

Having Type 2 diabetes or chronic kidney disease was associated with increased odds of COVID-19 related hospitalisation. Obesity was associated with an increased risk of admission to both hospital and ICU. People with dementia or those who had a stroke were twice as likely to die whereas they were less likely to end up in hospital or ICU, compared to people without.

These findings, highlighting the higher risk associated with clinical conditions in both isolation and in combination, are crucial for protecting these vulnerable cohorts as we are faced with the current surge and potential future waves of COVID-19 in the UK. Furthermore, identifying cohorts at increased risk of COVID-19 hospitalisation, ICU admission and death is also important for future vaccine planning as an effective vaccine becomes available.

Our study has some limitations. Firstly, in terms of data quality and availability, data on confirmed COVID-19 cases is highly likely to be an underestimation of the true number of cases and may also be skewed toward more severe cases given that. For example, testing was only done on people hospitalised with suspected COVID-19 up to 1 April 2020 with wider testing in the community available from May 2020 and capacity increasing throughout the year. Hence the data is limited to those who were recorded as having confirmed COVID-19 in primary or secondary care; and as such may omit many milder and asymptomatic cases that were unreported and untested.

Secondly, there were fewer deaths recorded in primary and secondary care data compared to the number of deaths reported by the ONS – 400 compared to 721 over the closest equivalent time period (18). Furthermore, the proportion of in-hospital deaths were disproportionately higher than the number of deaths taking place elsewhere. According to ONS data (19), 73% of COVID-19 deaths took place in hospital whereas 91% of deaths included in our analysis took place in a hospital. Also, it may be that a small number of COVID-19-related deaths were misclassified as non-COVID-19, particularly in the early stages of the pandemic when access to testing was limited. Accounting for this underestimation of mortality may alter our results. We also note that while primary care records are detailed and longitudinal, they may contain incomplete or out-of-date information.

Thirdly, 92% of the people included in the analysis live in an area that is within the 40% most deprived in England. As such, when IMD was included by quintile in the statistical analysis, the value of the regression coefficients was inconsistent or not statistically significant. Instead, a binary variable to indicate if people were living in the 30% most deprived areas nationally was included. We are in the process of collating more GP data from four other boroughs within NEL, which will allow us to expand our analysis and further investigate the impact of deprivation on hospitalisation, critical care and mortality outcomes in our future work.

Finally, we note that in our study we were unable to investigate the importance of additional factors such as household density, employment or access to personal protective equipment due to lack of access to such data. This is an important extension of our work that should be considered in future when such data becomes available.

Despite these limitations, our work is of notable importance as we identified new population cohorts that may benefit from protection during the current and future resurgences of COVID-19 in both NEL and more widely across the UK.

At the onset of the epidemic in the UK, shielding (staying at home at all times and avoiding any face-to-face contact) was recommended to protect extremely vulnerable groups. These recommendations were made based on pre-existing medical conditions, predominantly adapted from pre-existing recommendations for seasonal flu vaccination (20). While these recommendations have been crucial in protecting vulnerable groups to-date, updates must be made to guidance based on direct evidence of the risk factors for COVID-19 specifically (as opposed to related but distinct viruses), as more patient data becomes available. Using local data, we have been able to identify groups of people who are at increased risk of experiencing ill-health or dying following a diagnosis of COVID-19 – primarily those of older age, with multi-morbidity and those with learning disabilities but also those with dementia, previous stroke. These patient cohorts would benefit from preventative measures both in and out of hospital, through infection control and access to timely care.

It is also critical to consider those at increased risk of hospitalisation who survive COVID-19 and ensure they are supported in their recovery following hospital discharge. For example, people with Type 2 diabetes are at increased risk of a hospital admission (this may be due to COVID-19 or diabetes-related complications) whereas their risk of death is small. As such, tailored care pathways may need to be developed to support these groups to recover from COVID-19 and manage their pre-existing conditions following hospital discharge.

In summary, our study used established statistical methodology on linked primary and secondary data from a region at heightened risk of COVID-19 to confirm existing cohorts at risk of COVID-19 related hospitalisation, ICU admission and mortality. Overall, our work adds to the current body of evidence, drawing on locally linked primary and secondary care datasets to determine the key risk factors for COVID-19-related hospitalisation, ICU admission and mortality. This is crucially important as NEL, London and the UK face resurgent COVID-19 waves and planning is essential for managing demand and capacity and protecting vulnerable cohorts while awaiting a roll out of an effective vaccine.

Our findings have been essential in helping the North East London Integrated Health and Care System to develop equitable and appropriate epidemic response plans for managing COVID-19 locally as well as supporting the system and individuals recovering from COVID-19. Our work has important policy implications for existing and emerging public health initiatives to address wider determinants of health, and the work has been shared and discussed with scientific advisory bodies within the UK.

## Data Availability

All data and numerical code used in this analysis are available from the corresponding author upon reasonable request.

## Declarations

### Ethics approval and consent to participate

This study is considered a retrospective service evaluation and as such is exempt and ethics approval and informed consent for current study is waived by the UCL Research Ethics Committee.

### Availability of data and materials

The datasets used and analysed during this study and the numerical codes used to generate the outcomes of this paper are available from the corresponding author on reasonable request.

### Competing interests

We declare no competing interests.

### Patient and public involvement statement

Patients or the public were not involved in the design, or conduct, or report, or dissemination plans of this research.

## Notes

### Competing Interest Statement

The authors have declared no competing interest.

### Funding Statement

JPG acknowledges funding from NHS North East London Commissioning Alliance for this study.

## References

1. European Centre for Disease Prevention and Control. COVID-19 situation update world- wide, as of 17 September 2020 [Internet]. 2020 [cited 2020 Nov 4]. Available from: https://www.ecdc.europa.eu/en/geographical-distribution-2019-ncov-cases

2. UK Government. Coronavirus in the UK [Internet]. 2020 [cited 2020 Nov 5]. Available from: https://coronavirus.data.gov.uk/

3. Williamson EJ, Walker AJ, Bhaskaran K, Bacon S, Bates C, Morton CE, et al. Factors associated with COVID-19-related death using OpenSAFELY. Nature. 2020 Aug 1;584(7821):430–6.

4. Banerjee A, Pasea L, Harris S, Gonzalez-Izquierdo A, Torralbo A, Shallcross L, et al. Estimating excess 1-year mortality associated with the COVID-19 pandemic according to underlying conditions and age: a population-based cohort study. The Lancet. 2020 May 30;395(10238):1715–25.

5. Hull SA, Williams C, Ashworth M, Carvalho C, Boomla K. Suspected COVID-19 in primary care: how GP records contribute to understanding differences in prevalence by ethnicity. medRxiv. 2020 Jan 1;2020.05.23.20101741.

6. ONS. Deaths involving COVID-19 by local area and socioeconomic deprivation: deaths occurring between 1 March and 31 July 2020 [Internet]. 2020 [cited 2020 Jun 1]. Avail- able from: https://www.ons.gov.uk/peoplepopulationandcommunity/birthsdeathsandmarriages/deaths/bulletins/deathsinvolvingcovid19bylocalareasanddeprivation/deathsoccurringbetween1marchand31july2020

7. Rod JE, Oviedo-Trespalacios O, Cortes-Ramirez J. A brief-review of the risk factors for covid-19 severity. Revista de SaÃ °de PÃ °blica [Internet]. 2020;54. Available from: http://www.scielo.br/scielo.php?script=sci_arttext&pid=S0034-89102020000100701&nrm=iso

8. Deng G, Yin M, Chen X, Zeng F. Clinical determinants for fatality of 44,672 patients with COVID-19. Critical Care. 2020 Apr 28;24(1):179.

9. Docherty AB, Harrison EM, Green CA, Hardwick HE, Pius R, Norman L, et al. Features of 16,749 hospitalised UK patients with COVID-19 using the ISARIC WHO Clinical Characterisation Protocol. medRxiv. 2020 Jan 1;2020.04.23.20076042.

10. Lighter J, Phillips M, Hochman S, Sterling S, Johnson D, Francois F, et al. Obesity in Patients Younger Than 60 Years Is a Risk Factor for COVID-19 Hospital Admission. Clin Infect Dis. 2020 Jul 28;71(15):896–7.

11. ONS. ONS 2019 mid-year population estimates [Internet]. 2020 [cited 2020 Sep 20]. Available from: https://www.ons.gov.uk/peoplepopulationandcommunity/populationandmigration/populationestimates/bulletins/annualmidyearpopulationestimates/mid2019estimates

12. ONS via London Datastore. Ethnic Groups by Borough [Internet]. 2019 [cited 2020 Oct 10]. Available from: https://data.london.gov.uk/dataset/ethnic-groups-borough

13. UK Government. English indices of deprivation 2019 [Internet]. 2019 [cited 2020 May 1]. Available from: https://www.gov.uk/government/statistics/english-indices-of-deprivation-2019

14. Trust for London. Life expectancy by London borough, 2016-18 [Internet]. 2020. Avail- able from: https://www.trustforlondon.org.uk/data/life-expectancy-borough/

15. UK Government. New National Restrictions from 5 November [Internet]. 2020 [cited 2020 Nov 5]. Available from: https://www.gov.uk/guidance/new-national-restrictions-from-5-november

16. Commissioning Data Sets [Internet]. 2019 [cited 2020 Nov 2]. Available from: https://digital.nhs.uk/data-and-information/data-collections-and-data-sets/data-sets/commissioning-data-sets

17. Quality and Outcomes Framework [Internet]. 2019 [cited 2020 Jun 1]. Available from: https://digital.nhs.uk/data-and-information/publications/statistical/quality-and-outcomes-framework-achievement-prevalence-and-exceptions-data

18. ONS. Deaths involving COVID-19 by local area and deprivation [Internet]. 2020 [cited 2020 Sep 8]. Available from: https://www.ons.gov.uk/peoplepopulationandcommunity/birthsdeathsandmarriages/deaths/datasets/deathsinvolvingcovid19bylocalareaanddeprivation

19. ONS. Death registrations and occurrences by local authority and health board [Internet]. [cited 2020 Sep 8]. Available from: https://www.ons.gov.uk/peoplepopulationandcommunity/healthandsocialcare/causesofdeath/datasets/deathregistrationsandoccurrencesbylocalauthorityandhealthboard

20. NHSEI. Who’s at higher risk from coronavirus? [Internet]. 2020 [cited 2020 Oct 19]. Available from: https://www.nhs.uk/conditions/coronavirus-covid-19/people-at-higher-risk/whos-at-higher-risk-from-coronavirus/

